# Incorporating Preprints in Systematic Reviews: A Preliminary Study of a Novel Method for Rapid Evidence Synthesis

**DOI:** 10.1101/2025.07.15.25331581

**Authors:** Jiayi Tong, Yifei Sun, Rebecca A. Hubbard, M. Elle Saine, Hua Xu, Xu Zuo, Lifeng Lin, Chunhua Weng, Christopher Schmid, Stephen E. Kimmel, Craig A. Umscheid, Adam Cuker, Yong Chen

## Abstract

**Objectives:** By October 1, 2024, over 450,000 COVID-19 manuscripts were published, with 10% posted as unreviewed preprints. While they accelerate knowledge sharing, their inconsistent quality complicates systematic studies.

**Materials and Methods:** We propose a two-stage method to include preprints in meta-analyses. In Stage A, preprints are integrated through restriction or imputation and weighted by a confidence score reflecting their publication likelihood. In Stage B, we assess and adjust for potential publication or reporting biases.

**Results:** This preliminary study employed a two-stage procedure validated with two COVID-19 treatment case studies. For hydroxychloroquine, the relative risk (RR) was 1.06 [95% CI: 0.62, 1.80], suggesting no mortality benefit over placebo. For corticosteroids, the RR was 0.88 [95% CI: 0.62, 1.27], which, while not statistically significant, aligns with evidence supporting a mortality benefit.

**Discussion:** Our research aims to bridge a significant methodological gap by providing a solution for timely evidence synthesis, particularly in the face of the overwhelming number of publications surrounding COVID-19.

**Conclusion:** This preliminary study presents a method to efficiently synthesize COVID-19 research, including non-peer-reviewed preprints, to support clinical and policy decisions amidst the information surge.

## INTRODUCTION

As of October 1, 2024, over 700 million cases of SARS-CoV-2 and 7 million deaths have been recorded globally^1^. In response to the urgent demand for evidence-based treatment strategies, research findings on COVID-19 treatment effectiveness have proliferated since the pandemic began. By October 1, 2024, there were over 450,000 COVID-19 manuscripts available on PubMed and preprint platforms like bioRxiv and medRxiv ^2^. The National Library of Medicine (NLM) of the National Institutes of Health (NIH) has also made NIH-funded preprints accessible through PubMed Central (PMC) and, subsequently, PubMed. From June 2020 to January 2022, the NLM added over 3,500 preprints on NIH-backed COVID-19 studies to PMC and PubMed, with this number surging to more than 30,000 by October 01, 2024, when using the “preprint[filter]” search term. This vast influx of data poses challenges for decision-makers^3^. The scientific community is navigating not just the pandemic, but also an ’infodemic’—an overwhelming flood of publications^4^. Thus, the need for prompt and trustworthy evidence synthesis has never been more critical.

Among these systematic reviews, there are more than 100 living systematic reviews, which are continually updated based on new emerging evidence. The continuously updating feature of a living systematic review improves the validity of conclusions. It also aids readers in keeping pace with a fast-moving field by providing an up-to-date summary of the evidence. This is particularly relevant for meta-analyses (which use statistical methods to quantitatively synthesize evidence from multiple studies to achieve a generalizable and reliable pooled estimate) and cumulative meta-analyses (which are meta-analyses that are updated as new evidence appears for temporal trends of intervention effects). For investigating COVID-19 treatment effectiveness, many meta-analyses have been conducted. These studies offer researchers and clinicians information to better understand intervention effectiveness for patients infected with COVID-19.

Systematic reviews stand as the gold standard in collating empirical evidence from diverse studies, offering the highest level of evidence for scientific questions. As of now, PROSPERO, an international registry for systematic reviews, has registered over 18,000 COVID-19-specific protocols^5^. Within this collection, over 100 are living systematic reviews—reviews that undergo continuous updates with emerging evidence^6^. This dynamic updating enriches the validity of the conclusions and equips readers with current summaries in rapidly evolving fields^7^. This practice is especially critical for meta-analyses, which pool evidence from multiple studies for a consistent and reliable estimate^8^, and for cumulative meta-analyses that track the temporal trends of intervention impacts^9^. As an example, numerous meta-analyses investigating COVID-19 treatment effectiveness have been undertaken^10–12^, providing invaluable insights into intervention efficacy for COVID-19 patients.

While systematic reviews promise high-quality and timely evidence synthesis, COVID-19 introduces unique challenges. One of the key challenges is the inclusion of data from preprint servers, such as bioRxiv and medRxiv, which now account for a substantial portion of available evidence. The use of preprint servers in the life sciences has surged recently. These platforms have been pivotal in real-time understanding and response to COVID-19, bypassing the usual publication cycle delays. In meta-analyses, including preprints can offer immediate and impactful findings. Notably, the RECOVERY Collaborative Group shared pivotal results on medRxiv about the efficacy of dexamethasone for hospitalized COVID-19 patients—a study later peer-reviewed and published after 248 days ^13^. Other notable preprints cover topics like hydroxychloroquine’s preventive potential for COVID-19 and the effects of convalescent plasma treatment on severe cases^13–16^. However, many meta-analyses blend preprints with peer-reviewed studies, overlooking the differing probabilities of formal publication^10,11^. Given that not all preprints endure the peer-review process, unselective inclusion of all preprints can bias conclusions and produce misleading clinical guidance. Hence, the likelihood of a preprint’s eventual publication should be a factor in the process of evidence synthesis^2,17^. Studies with higher publication potential should be given greater weights over those with lower probability of being published.

The second significant challenge is the influence of publication or reporting bias, arguably the most critical concern for the credibility of meta-analyses^18^. This bias manifests when the publication of a manuscript’s results depends on their significance or direction. Given today’s research landscape and political environment^19^, the likelihood of such bias in both published papers and preprints is especially high for COVID-19 studies. Systematic reviews and meta-analyses must, therefore, factor in the potential impacts of publication or reporting bias.

Driven by the pressing need to address these challenges, our objective in this paper is to design and validate a two-stage method for evidence synthesis. This approach integrates preprints using a rigorous procedure that concurrently accounts for both their publishability (the likelihood a preprint will be published) and publication bias. This refined two-stage method promotes the appropriate inclusion of up-to-date findings from preprints, fostering more prompt and trustworthy conclusions in meta-analyses with reduced publication bias.

## Methods and Materials

### Data

We collected 146 preprints on COVID-19 treatments from bioRxiv and medRxiv through 09/03/2020 and manually extracted 10 features suggested by our clinical team (ES, SK, CU and AC), including study type (i.e., RCT, observational study, others), median age of patients included in the study, sample size, single or multi-center study design, whether or not the results were preliminary, whether or not the analysis was adjusted for confounding variables, country of study, citation counts in first two weeks, number of PDFs downloaded from preprint servers in first two weeks, and h-index of the last author from Google scholar on 09/03/2020. These features were selected to capture early signals of downstream impact, combining study-level metadata (e.g., age, country) with early dissemination metrics (e.g., downloads). While some may be subjective, they reflect both content and engagement factors relevant to systematic review inclusion. The same feature set was used across both case studies. More details on data resources are provided in Supplementary Appendix A.

### A novel two-stage approach advances meta-analysis compared with traditional methods

In Figure 1, we present an illustration of the proposed two-stage method. Traditional meta-analyses typically use the following two strategies: (I) including published studies only; or (II) combining published studies and all preprints. Different from the traditional meta-analysis strategy, our proposed approach incorporates preprints by considering the publication probability, which we referred to as the confidence score^20^ (CS): (III) using CS = 0·5 as a cutoff value for the preprint inclusion. If CS < 0·5, the preprint is not included; if CS is >= 0·5, the preprint is included as a published study; or (IV) including all preprints, and using CS to weight preprints with the multiple imputation method). In addition, after incorporating preprints using CS, we performed a sensitivity analysis for publication bias (Stage B. V in Figure 1). In particular, strategy (III) offers simplicity and interpretability, allowing for a binary inclusion/exclusion rule based on a confidence score. However, this may result in the loss of potentially useful information from lower-scoring preprints. On the other hand, strategy (IV) retains the full spectrum of preprints and adjusts their influence proportionally to their estimated likelihood of publication. This approach is more inclusive and statistically principled. More details of strategy (IV) are introduced below. Further discussion on the choice between strategies (III) and (IV) can be found in Supplementary Appendix Section D.

**Figure 1.**
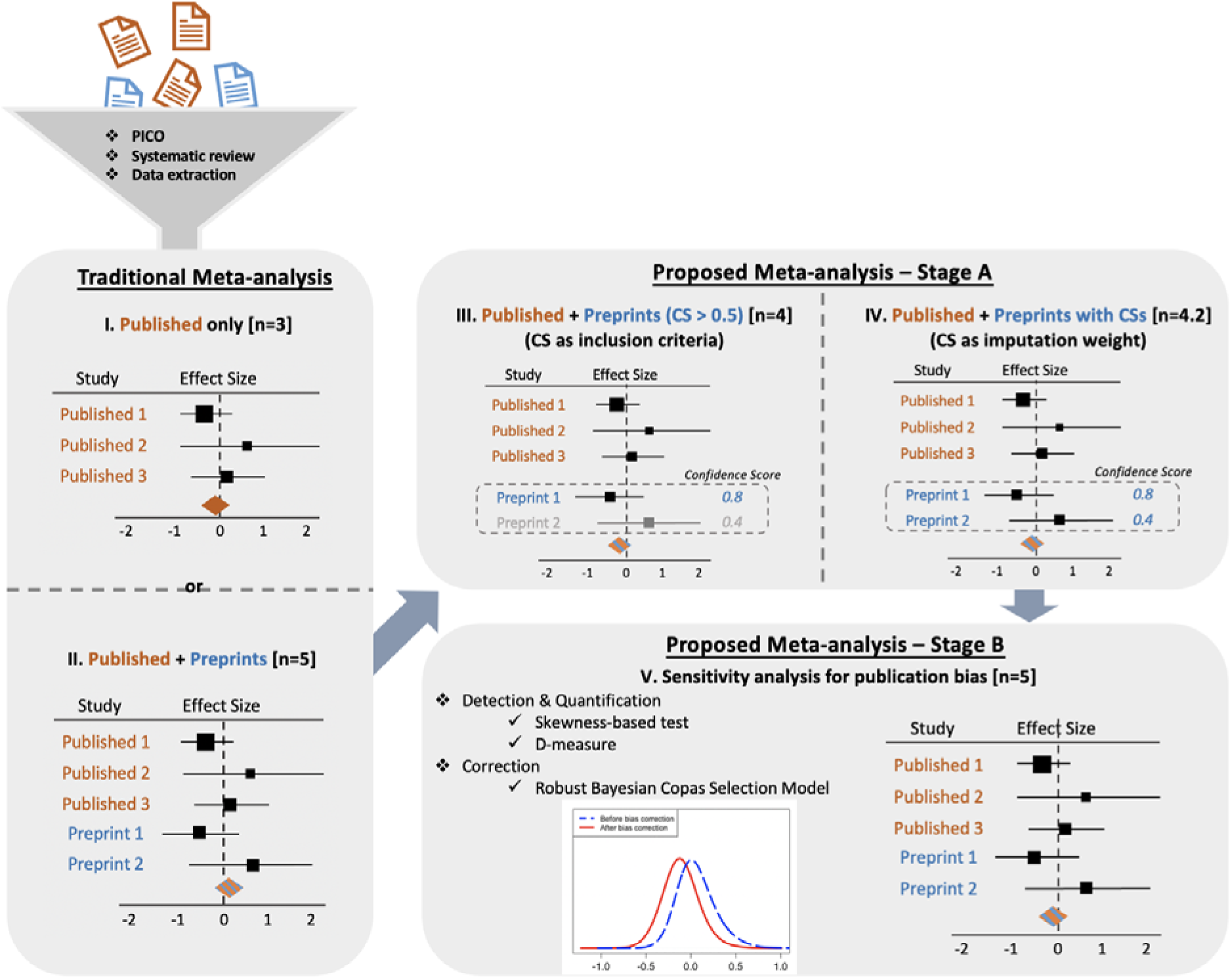
An illustration of the pipeline for the proposed two-stage meta-analysis approach. In the traditional meta-analysis panel on the left, two commonly used meta-analysis strategies are presented: (I) including published articles only and (II) combining published articles and all preprints. For the proposed meta-analysis methods in the right panel, the two-stage procedure accounts for the heterogeneous publication probability of preprints (III, IV), while simultaneously addressing potential publication/reporting bias (V). The size of the black box in each forest plot is proportional to the sample size of the corresponding study. [CS: Confidence score; PICO: Population, Intervention, Comparison, Outcome]

### Stage A: Predictive model and confidence score (CS)

To predict the probability that a preprint will be published, we used a survival mixture model^21^. Cure models are typically used in cases where the survival distribution is believed to arise from a mixture of patients who will never experience the outcome of interest (i.e., are “cured”) and those who will experience the outcome. In our context, we consider publication to be the outcome of interest, so preprints that will never be published are considered “cured” (i.e., never experience the outcome of interest, which is publication). We also evaluate the time from posting on preprint servers to publication in a peer-reviewed journal. Given our primary goal of predicting the probability of a preprint being published, the rationale for using a survival mixture model is based on the assumption that preprints can be viewed as a mixture of two groups: those that will never be published and those with a non-zero probability of eventual publication. Since the timing of preprint posting is a critical factor, the survival mixture model enables us to capture the time from posting to either publication or administrative censoring. This framework reflects the intuition that preprints remaining on servers for a longer duration (e.g., several years) are less likely to be published compared to those that have been posted more recently (e.g., within a few days or weeks). The publication time is subject to administrative censoring due to data freeze on 09/03/2020. More details the method are available in the Supplementary Materials Appendix Section B.

### Stage B: Quantification and Correction of Publication/Reporting Bias

Several statistical methods exist to mitigate publication/reporting bias, such as the commonly used funnel plot^22^, Egger’s test^23^, Copas model^24,25^, and skewness based measure^26^. In this work, we introduced a novel Bayesian Copas Selection model that addresses publication/reporting bias. Our method refines the implementation of Copas selection model which are typically difficult to fit^27,28^, and introduces a novel measure, named as D-measure, for quantifying publication bias based on the Hellinger distance between effect size distributions pre and post bias adjustment. This measure provides clear interpretation as detailed in Table 1. Intuitively, the D-measure captures how much the estimated results change after accounting for possible publication/reporting bias. If the results barely change, the D-measure is close to zero, suggesting little evidence of bias. On the other hand, a large D-measure (i.e., closer to one) means the effect size distribution has shifted considerably after adjustment, indicating a high likelihood that publication bias influenced the original findings. Such quantity provides a simple, interpretable number to assess the extent of publication/reporting bias in a meta-analysis.

**Table 1.** Interpretation of the D-measure. D-measure is based on the Hellinger distance between the posterior distributions of the effect size before and after bias correction. It directly quantifies the impact of publication bias on the inference on effect size and considers the entire posterior distribution. D-measure takes values between 0 and 1, with 0 being no difference in posterior densities (i.e., no publication bias) and 1 being vastly different (i.e., substantial publication bias).

| Range | Interpretation |
| --- | --- |
| $0.00 \leq D \leq 0.25$ | Negligible Publication Bias |
| $0.25 \leq D \leq 0.50$ | Moderate Publication Bias |
| $0.50 \leq D \leq 0.75$ | High Publication Bias |
| $0.75 \leq D \leq 1.00$ | Very High Publication Bias |

### Patients and Public Involvement

There were no patients or the public involved in the design, or conduct, or reporting, or dissemination plans of our research.

## Results

### Validation of Confidence Scores for Predicting the Likelihood of Publication of Preprints

Of the 146 preprints, 23 (16%) were published by 09/03/2020. Using the 10 extracted features, the cure model produced confidence scores (probabilities of a preprint’s publication) for both unpublished and published preprints. Figure 2 illustrates that the median scores are 0.001 for the yet-to-be-published preprints and 0.997 for those published. These scores, derived from the survival mixture model, effectively distinguish between the two groups. Given the dataset’s size, we employed leave-one-out cross-validation instead of data splitting. The resultant AUC was 0.89 (95% CI: 0.81, 0.97).

**Figure 2.**
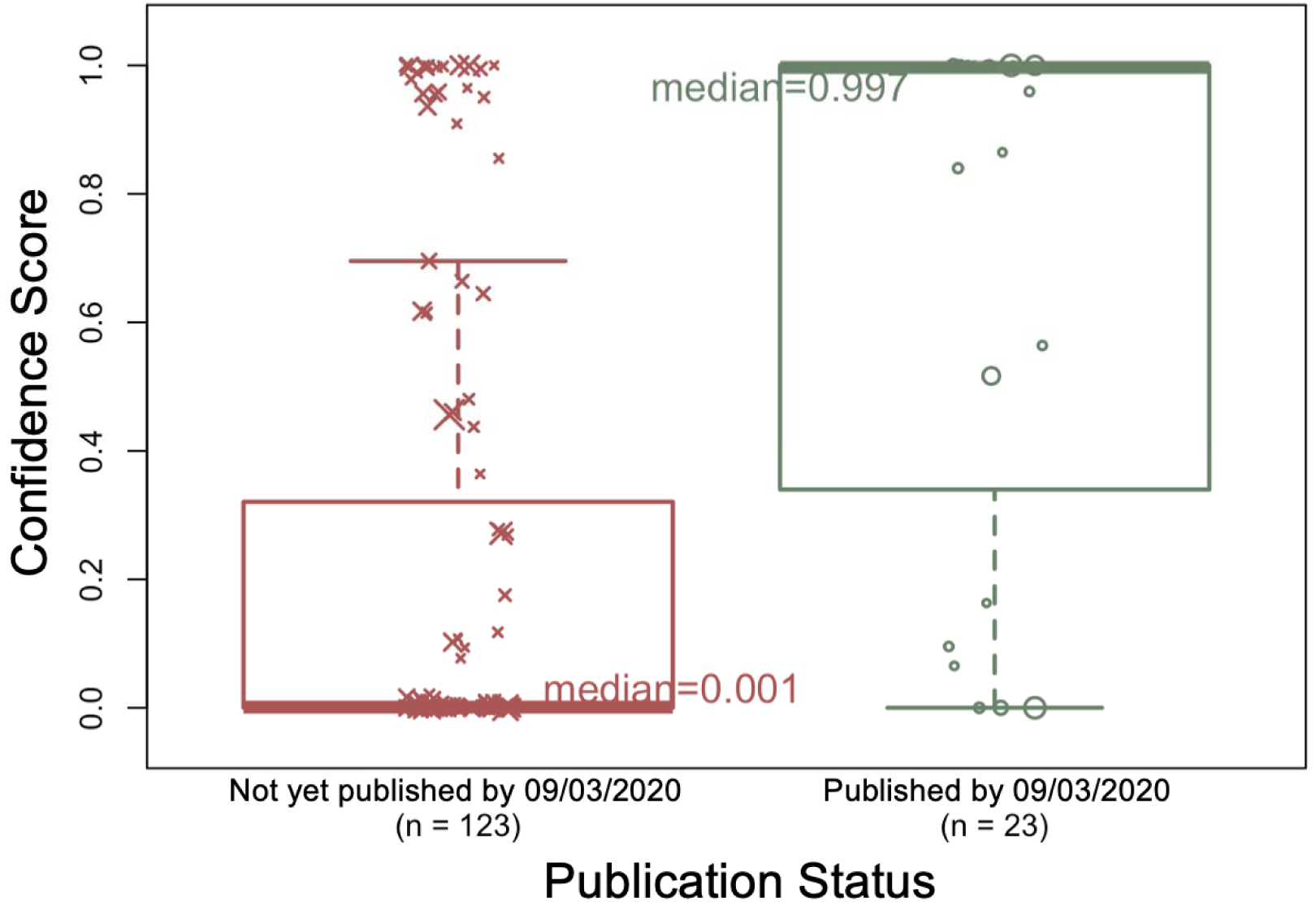
Comparison of the confidence scores for the unpublished by 09/03/2020 preprints (left box [red]) and th published preprints (right box [green]). As of 09/03/2020, 123 of the 146 preprints were not yet published and 84 of them were published. The median confidence score of the not-yet-published preprints was 0, which is more than 99% lower than the median confidence score of the published preprints (0.997) (p-value = 3.24e-10 << 0·01).

### Case study 1 -- Hydroxychloroquine

Early in the pandemic, on March 28, 2020, hydroxychloroquine received an emergency use authorization (EUA) from the FDA to treat hospitalized COVID-19 patients. However, an increasing number of studies have shown that hydroxychloroquine provides no mortality benefit to patients with COVID-19. The NIH halted clinical investigations on hydroxychloroquine in mid-2020, and the EUA was revoked on June 15, 2020^29^. The ineffectiveness of hydroxychloroquine in COVID-19 provides a useful test case because the relative risk between the treatment group and control group (i.e., placebo or standard care) is likely to be close to 1^30^.

We compared four meta-analysis strategies in this case study. We identified 6 published articles from PubMed, of which the meta-analysis estimated RR of mortality was 1.49 (95% confidence interval [CI]: 1.04, 2.16), suggesting harm from the medication. We identified 7 preprints examining the effectiveness of hydroxychloroquine that had not been published as of the censoring date from the 146 preprints screened. The PRISMA diagram is available in the Supplementary Appendix Section A. By simply combining the 7 preprints and 6 published papers using traditional meta-analysis, the RR was 1.37 (95% CI: 0.84, 2.25). With strategy (III), 4 preprints with CSs larger than 0·5 were combined with the published articles using meta-analysis. The RR with this approach was 1.02 (95% CI: 0.68, 1.54). In strategy (IV), we used the CS to impute a publication status for each preprint. The RR of this meta-analysis was 1.02 (95% CI: 0.67, 1.54). By properly including preprints (i.e., either using a cutoff based on estimated CS, or weighted by CS), our analyses have led to estimated RRs close to 1 (Figure 3), which is consistent with the emerging clinical consensus and existing studies and meta-analyses indicating that hydroxychloroquine is ineffective^31–34^. For example, Kashour et al. analyzed thirteen studies (one randomized controlled trial and twelve cohort studies, where three studies were preprints) involving 15,938 hospitalized patients to evaluate the effect of hydroxychloroquine on short-term mortality. The pooled adjusted odds ratio was 1.05 (95% CI: 0.96–1.15)^34^.

**Figure 3.**
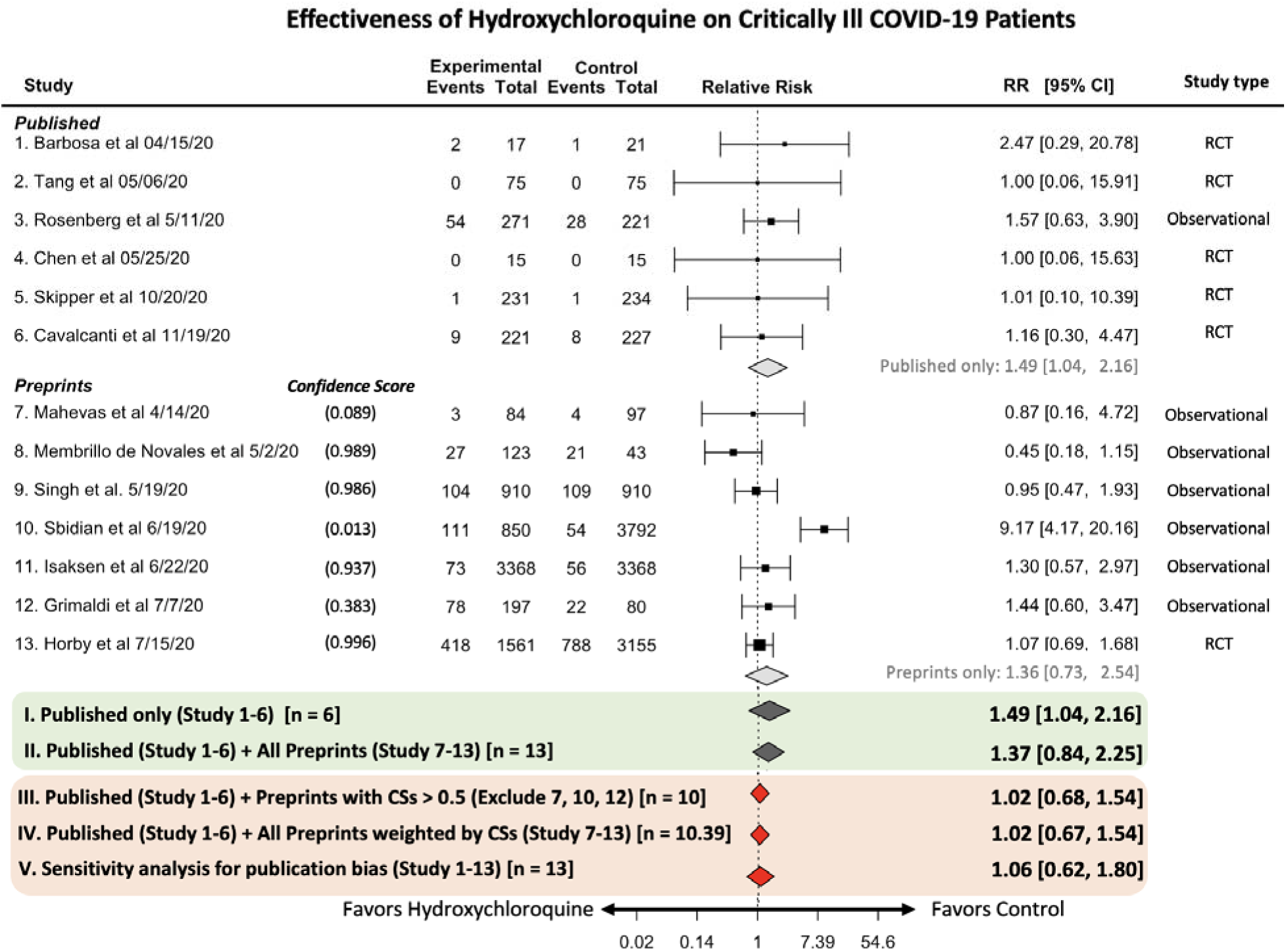
Forest plot of the association between hydroxychloroquine and mortality in critically ill COVID-19 patients and a comparison between four meta-analysis strategies. Studies 1-6 are published and studies 7-13 are preprints that were not published as of 09/03/2020. Within the green square are the two most used traditional meta- analysis strategies. In the orange square, (III) and (IV) are the strategies that use confidence scores. Compared with strategies (I) and (II), (III) and (IV) have smaller relative risks that are closer to 1. Strategy (V) is the sensitivity analysis for publication bias. Strategy (V) includes a publication bias correction on top of (IV) and gives the most reliable relative risk.

In Figure 3, we present the forest plot and relative risk for the four different meta-analysis strategies. Using the confidence scores either as an inclusion criterion or imputation weight in the meta-analysis, the relative risk was closer to unity compared with ignoring or simply combining preprints (i.e., strategies (I) and (II)).

In Stage A, both strategies (III) and (IV) used the confidence score to incorporate preprints into the meta-analysis. In strategy (IV), no preprints were excluded. All evidence in preprints was synthesized in the weighted meta-analysis. Thus, on top of strategy (IV), we further conducted a sensitivity analysis for publication bias as Stage B of the proposed procedure. We quantified and corrected the potential publication bias. We applied the skewness-based test and the D- measure to quantify publication bias after carrying out strategy (IV). If publication bias was detected with a p-value of the skewness-based test smaller than 0.05 or D-measure larger than 0.25, we further applied the robust Bayesian Copas Selection Model to correct the publication bias.

The p-value of the skewness test on strategy (IV) from Stage A was 0.84. The D-measure was 0.04, which falls into the smallest category (Table 1). Based on both measures, the publication/reporting bias of the papers and preprints on hydroxychloroquine is negligible. This small bias indicates that both preprints and peer-reviewed papers were not selective in posting or publication with respect to the magnitude of the estimated RR.

We further quantify the reporting bias for preprints only and publication bias for the published papers only. For preprints only, the p-value of the skewness test was 0.47. The value of the D- measure was 0.04, indicating that publication/reporting bias is negligible. For the published studies only, the p-value was 0.57 and the D-measure was 0.05. Both indicate that there was no publication/reporting bias for the published papers. These results show that for both preprints and published papers on hydroxychloroquine, the data do not suggest the presence of reporting/publication bias. With negligible bias, the correction step is optional. However, for completeness, we corrected for publication bias, and the relative risk after bias correction was 1.06 (95% CI: 0.62, 1.80) (Table 2), indicating that there is evidence showing that that hydroxychloroquine is ineffective in reducing the death rate for the clinically ill patient infected by COVID-19.

**Table 2.**
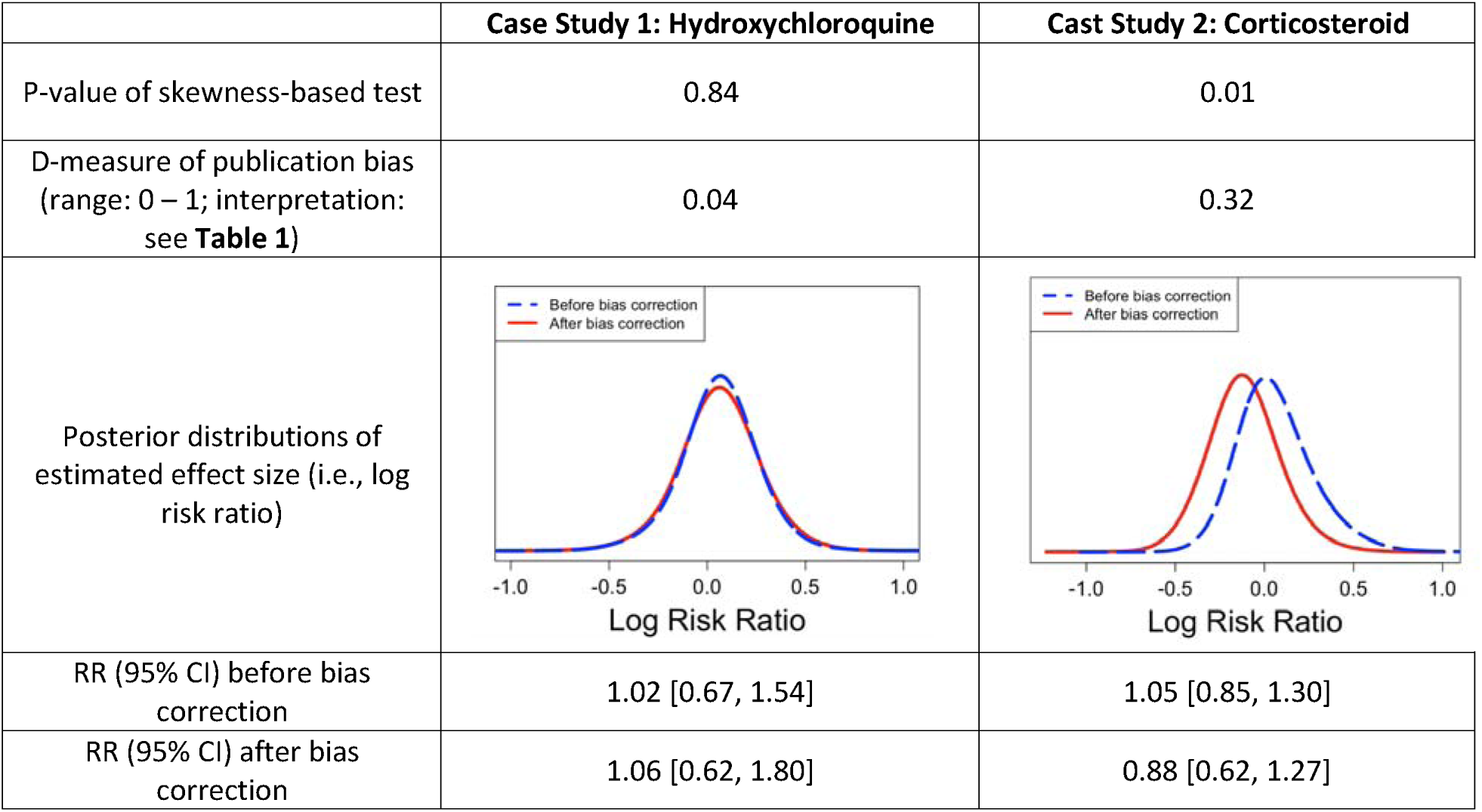
Results of the quantification and correction of publication bias. For case study 1, the large p-value and small D-measure value provide no evidence of publication bias, but for completeness, we also correceted for pubcaltion bias. The posterior distributions of the estimated log risk ratio are close, leading to relative risks (RR) of 1.02 before and 1.06 after bias correction. For case study 2, the small p-value and moderate D-measure value indicate the presence of publication bias. The posterior distribution of the estimated log risk ratio after bias correction is shifted towards the left. The RR after bias correction is 0.88 with 95% CI (0.62, 1.27), suggesting that corticosteroids may reduce the mortality rate for critically ill COVID-19 patients.

|  | Case Study 1: Hydroxychloroquine | Case Study 2: Corticosteroid |
| --- | --- | --- |
| P-value of skewness-based test | 0.84 | 0.01 |
| D-measure of publication bias<br>(range: 0 – 1; interpretation:<br>see Table 1) | 0.04 | 0.32 |
| Posterior distributions of<br>estimated effect size (i.e., log<br>risk ratio) |  |  |
| RR (95% CI) before bias<br>correction | 1.02 [0.67, 1.54] | 1.05 [0.85, 1.30] |
| RR (95% CI) after bias<br>correction | 1.06 [0.62, 1.80] | 0.88 [0.62, 1.27] |

### Case study 2 -- Corticosteroids

The second case study is a meta-analysis of the effectiveness of corticosteroids. Recent meta-analyses showed that corticosteroids are likely to reduce mortality in severe COVID-19 patients compared with usual care or placebo^10,11^. However, the opposite conclusion in other existing meta-analyses of observational studies^35^ (i.e., that use of corticosteroids delayed recovery and did not improve mortality for critically ill patients.) motivated us to conduct a rigorous meta-analysis with the proposed two-stage method.

In this case study, we compared four meta-analysis strategies. The relative risk (RR) for mortality in the 19 published peer-reviewed papers from PubMed was 1.10 (95% CI: 0.88, 1.38). By simply combining the 7 eligible preprints and 19 peer-reviewed papers using traditional meta-analysis, we obtained a relative risk of 1.01 (95% CI: 0.84, 1.22). With a threshold of 0·5 for the confidence scores for inclusion in the meta-analysis, we included 3 out of the 7 preprints in the meta-analysis and the risk ratio was 1.03 [95% CI: 0.83, 1.27]. When using confidence scores as imputation weights for the preprints, the RR was 1.05 (95% CI: 0.85, 1.30) (Figure 4). Use of confidence scores in strategy (III) and (IV) down-weighted the preprints (RR for preprints only: 0.76, 95% CI: 0.48, 1.22) compared with strategy (II).

**Figure 4.**
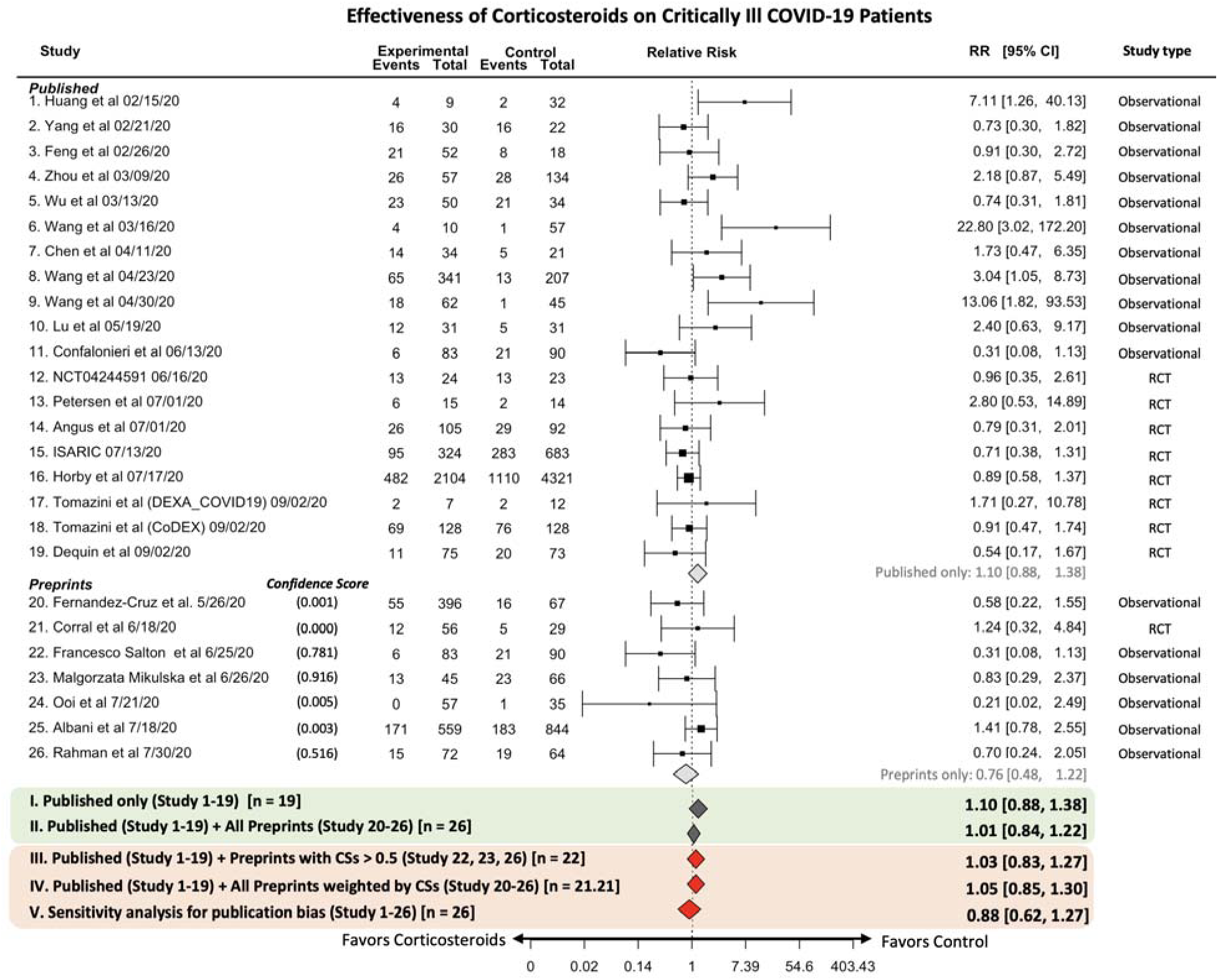
Forest plot of association between corticosteroids and mortality in critically ill COVID-19 patients and comparison between four meta-analysis strategies. Studies 1-19 are published and studies 20-26 are preprints that had not been published as of 09/03/2020. Within the green square are the two most used traditional meta- analysis strategies. In the orange square, strategies (III) and (IV) use confidence scores. Strategy (V) is th sensitivity analysis for publication bias. Strategy (V) performs the publication bias correction on top of (IV) and gives the most reliable relative risk.

From the above results, we also observe that in both case studies 1 and 2 there is a single RCT included among the preprints. The confidence score for the preprint RCT is relatively large in case study 1 (study 13, CS = 0.996) but is extremely small in case study 2 (study 21, CS = 0.000). Although common intuition suggests that an RCT is more likely to be published than an observational study, the RCT design alone cannot guarantee publication. In the predictive model, other factors such as length of time the paper has been posted on the preprint server, sample size, citation count within the first two weeks posted on preprint servers, h-index of the last author, and others also contribute to the probability of publication as estimated by the confidence score.

We repeated similar steps as in case study 1. After applying strategy (IV), the p-value of the skewness test was 0·01. The D-measure was 0·34, which represents moderate publication bias. Both results indicate the existence of publication bias among the 26 studies on corticosteroids. Thus, we further corrected the bias with the robust Bayesian Copas Selection model. After this correction, the relative risk for mortality was 0.88 (95% CI: 0.62, 1.27). Though not statistically significant, the direction of effect is consistent with the body of evidence and meta-analysis studies that has emerged suggesting that corticosteroids are beneficial in reducing COVID-19 mortality^36,37^. After bias correction (solid red line), the posterior distribution of the log risk ratio (Table 2) moved toward the left and is clearly separated from the posterior distribution before bias correction (dashed blue line).

Furthermore, we investigated publication/reporting bias for published papers only and preprints only. The p-value of the skewness test for preprints only was 0.09, and the D-measure for preprints only was 0.07. Based on these results, there is a lack of evidence suggesting the presence of reporting bias for preprints. For the published studies only, the p-value is 0.01 and the D-measure was 0.32, suggesting the existence of publication bias of the published papers examining the effectiveness of the corticosteroids for COVID-19 patients.

Sub-analysis results with RCTs only for both use cases are provided in Supplementary Appendix C.

## DISCUSSION

In this study, we introduced a two-stage procedure to address two pivotal challenges: accounting for publication probabilities of preprints and counteracting publication/reporting bias in the synthesis of evidence related to COVID-19 from both peer-reviewed articles and preprints. Our approach promotes the timely and trustworthy incorporation of preprint findings into meta-analyses, ensuring conclusions minimally impacted by publication/reporting bias. Through case studies on COVID-19 treatments, namely hydroxychloroquine and corticosteroids, our results aligned with primary research on these treatments.

Our methodology offers immense potential to invigorate living systematic reviews, especially in fast-paced domains like the formulation of clinical practice guidelines. The ongoing exploration of COVID-19 treatments expects a growing number of preprints. Integrating these preprints through continuous monitoring in living systematic reviews ensures the persistent validity and relevance of synthesized findings ^31,32^ ^10,11,38^. Importantly, our method underscores that preprints, being repositories of timely information, shouldn’t be sidelined in meta-analyses. Yet, if they are considered, their potential for eventual publication should be a pivotal consideration. This strategy offers a timely inclusion approach, particularly fitting for rapidly evolving topics.

The proposed framework is designed to serve as a complementary tool for evidence synthesis when incorporating preprints in meta-analyses. This framework allows researchers to assess how inclusion of preprints, weighted by their likelihood of eventual publication, might influence overall conclusions. Within this framework, we offer two strategies: applying a threshold on the confidence score, i.e., strategy (III), or weighting all preprints by their confidence scores, i.e., strategy (IC). While both strategies yielded similar results in our case studies, they serve different analytical needs. Strategy (III) is straightforward and easier to implement, making it suitable when researchers prefer a binary inclusion rule. However, it may exclude informative preprints with moderately high scores. In contrast, strategy (IV) offers a more continuous and precise approach, allowing all preprints to contribute to the synthesis in proportion to their estimated publication probability. Ultimately, the choice between these strategies depends on the context and goals of the review. For example, users seeking interpretability and simplicity may favor thresholding, while those emphasizing inclusiveness may prefer weighting. Our results suggest that either approach, when coupled with publication bias correction, can enhance the credibility of meta-analytic findings involving preprints.

This study represents a preliminary but important step toward developing a scalable two-stage framework for incorporating preprints into real-world evidence synthesis. While the results are promising, we acknowledge that further external validation is necessary to assess the robustness and generalizability of the proposed approach across broader datasets and settings. First of all. while our model accounts for the time from preprint posting to publication using a cure survival framework, we recognize that various underlying factors may influence this time-to-publication process. Although we have incorporated a set of key risk factors informed by domain knowledge and expertise, we acknowledge that these covariates may not capture the full range of relevant influences. Future research could enhance this framework by integrating additional predictors, such as editorial policies, journal scope, and evolving trends in scientific attention, while also addressing the subjectivity introduced by using citation counts in computing the confidence score. To ensure robustness and relevance, future work should incorporate a dedicated, data-driven feature selection step, such as regularized regression or ensemble-based importance ranking, complemented by clinical or domain-expert review. Moreover, given the heterogeneity across scientific disciplines, the optimal set of predictive features is likely to be domain-specific. Tailoring the feature set to reflect the norms, dissemination patterns, and evidence standards of each research area will be essential to improving the generalizability and interpretability of the framework.

Although our confidence score aims to complement, not replace, current study quality metrics, future endeavors might focus on incorporating a broader range of qualitative and quantitative measures, such as the Cochrane Risk of Bias (ROB) 2.0 Tool ^39^ ^39^for RCTs, Newcastle-Ottawa Scale (NOS) for nonrandomized studies, and CONSORT/STROBE compliance^40,41^. In our case studies, we chose to combine both RCTs and observational studies, which enhanced our sample size but might affect the accuracy of the derived efficacy estimates. We concentrated on treatment effectiveness, aiming to sidestep potential pitfalls linked with limited sample sizes. We emphasized all-cause mortality, suggesting future research might further stratify results based on different follow-up durations and study types. A key limitation of this study is the modest sample size of preprints used to train the predictive model, which may limit the generalizability of our findings. This limitation is partly driven by the static nature of our dataset, which captures publication status as of the censoring date (09/03/2020). While this snapshot allowed us to evaluate the framework under realistic conditions, it does not reflect the dynamic nature of publication activity—particularly relevant in the context of living systematic reviews. Expanding the dataset across other disease areas or longer time windows, and updating publication outcomes beyond the current cutoff, would improve both the robustness and applicability of our approach. However, the process of tracking publication status and extracting relevant metadata is currently manual and resource-intensive. As such, future work will focus on developing an automated pipeline for metadata extraction and publication tracking. This would significantly improve scalability, enable real-time integration of newly published studies, and enhance the framework’s utility for living evidence synthesis.

The effort to integrate preprints into broader academic discussions is still in its early stages. Future directions may involve refining the confidence score’s predictive model by integrating a more extensive predictor set and harnessing natural language processing (NLP) and large language models (LLM) such as ChatGPT to automate feature extraction. Enhanced collaboration across research entities will foster a deeper comprehension of our proposed methodology. As the domain evolves, continuous effort is crucial to delve deeper into the dynamic interplay between preprints, peer-reviewed research, and the overarching narrative of research dissemination.

## Data Availability

The data that support the findings of this study are available from the corresponding author upon request.

## Acknowledgement

We thank Anchita Batra, Anni Pan, and Oliva Wang from University of Pennsylvania, for their contribution in data collection and extraction for the case studies. These students were supported by 2020 Penn Undergraduate Research Mentoring Program (PURM). All of the analyses were conducted using R version 4.3.3.

## Author Contributions

JT and YC developed the methods; JT and MES collected the data from PubMed and preprints server; YC and AC guided the implementation of case studies; YC conceived the idea; JT and YC developed and implemented the methods; JT conducted the two case studies; All authors interpreted the results and provided instructive comments; JT and YC drafted the main manuscript. All authors have approved the manuscript.

## References

1. COVID-19 cases | WHO COVID-19 dashboard. https://data.who.int/dashboards/covid19/cases?n=c.

2. Fraser, N. et al. Preprinting the COVID-19 pandemic. bioRxiv 2020.05.22.111294 Preprint at 10.1101/2020.05.22.111294 (2020).

3. Coronavirus: How much news is too much? - BBC Worklife. https://www.bbc.com/worklife/article/20200505-coronavirus-how-much-news-is-too-much.

4. Orso, D., Federici, N., Copetti, R., Vetrugno, L. & Bove, T. Infodemic and the spread of fake news in the COVID-19-era. Eur J Emerg Med 27, 327–328 (2020).

5. PROSPERO. https://www.crd.york.ac.uk/prospero/#searchadvanced.

6. Maguire, A., Douglas, I., Smeeth, L. & Thompson, M. Determinants of cholesterol and triglycerides recording in patients treated with lipid lowering therapy in UK primary care. Pharmacoepidemiol Drug Saf 16, 228–228 (2007).

7. Elliott, J. H. et al. Living systematic review: 1. Introduction—the why, what, when, and how. J Clin Epidemiol 91, 23–30 (2017).

8. Ahn, E. & Kang, H. Introduction to systematic review and meta-analysis. Korean J Anesthesiol 71, 103–112 (2018).

9. Lau, J. et al. Cumulative meta-analysis of therapeutic trials for myocardial infarction. N. Engl. J. Med. 327, 248–254 (1992).

10. Siemieniuk, R. A. C., et al. Drug treatments for covid-19: Living systematic review and network meta-Analysis. The BMJ 370, (2020).

11. Sterne, J. A. C. et al. Association between Administration of Systemic Corticosteroids and Mortality among Critically Ill Patients with COVID-19: A Meta-analysis. JAMA - Journal of the American Medical Association 324, 1330–1341 (2020).

12. Axfors, C. et al. Mortality outcomes with hydroxychloroquine and chloroquine in COVID- 19 from an international collaborative meta-analysis of randomized trials. Nat Commun 12, 1–13 (2021).

13. Dexamethasone in Hospitalized Patients with Covid-19. New England Journal of Medicine 384, 693–704 (2021).

14. Tang, W. et al. Hydroxychloroquine in patients with mainly mild to moderate coronavirus disease 2019: open label, randomised controlled trial. bmj 369, (2020).

15. Mitjà, O. et al. A cluster-randomized trial of hydroxychloroquine for prevention of Covid-19. New England Journal of Medicine (2020).

16. Liu, S. T. H. et al. Convalescent plasma treatment of severe COVID-19: a propensity score–matched control study. Nat Med 26, 1708–1713 (2020).

17. Van Schalkwyk, M. C. I., Hird, T. R., Maani, N., Petticrew, M. & Gilmore, A. B. The perils of preprints. The BMJ vol. 370 Preprint at 10.1136/bmj.m3111 (2020).

18. Dickersin, K. Publication Bias: Recognizing the Problem, Understanding Its Origins and Scope, and Preventing Harm. in Publication Bias in Meta-Analysis 9–33 (John Wiley & Sons, Ltd, Chichester, UK, 2006). doi:10.1002/0470870168.ch2.

19. Berenbaum, M. R. On COVID-19, cognitive bias, and open access. Proceedings of the National Academy of Sciences of the United States of America vol. 118 Preprint at 10.1073/PNAS.2026319118 (2021).

20. Tong, J. et al. Confidence Score: A Data-Driven Measure for Inclusive Systematic Reviews Considering Unpublished Preprints. Journal of the American Medical Informatics Association (2023) doi:10.1093/JAMIA/OCAD248.

21. Othus, M., Barlogie, B., LeBlanc, M. L. & Crowley, J. J. Cure models as a useful statistical tool for analyzing survival. Clinical Cancer Research vol. 18 3731–3736 Preprint at 10.1158/1078-0432.CCR-11-2859 (2012).

22. Light, R. J. & Pillemer, D. B. Summing uprl: the science of reviewing research. 191 (1984).

23. Egger, M., Smith, G. D., Schneider, M. & Minder, C. Bias in meta-analysis detected by a simple, graphical test. BMJ 315, 629–634 (1997).

24. Copas, J. B. & Shi, J. Q. A sensitivity analysis for publication bias in systematic reviews. Stat Methods Med Res 10, 251–265 (2001).

25. J, C. & JQ, S. Meta-analysis, funnel plots and sensitivity analysis. Biostatistics 1, 247–262 (2000).

26. Lin, L. & Chu, H. Quantifying publication bias in meta-analysis. Biometrics 74, 785–794 (2018).

27. Ning, J., Chen, Y. & Piao, J. Maximum likelihood estimation and EM algorithm of Copas- like selection model for publication bias correction. Biostatistics 18, 495–504 (2017).

28. Chen, Y., Ning, J. & Cai, C. Regression analysis of longitudinal data with irregular and informative observation times. Biostatistics 16, 727–739 (2015).

29. Coronavirus (COVID-19) Update: FDA Revokes Emergency Use Authorization for Chloroquine and Hydroxychloroquine | FDA. https://www.fda.gov/news-events/press-announcements/coronavirus-covid-19-update-fda-revokes-emergency-use-authorization-chloroquine-and.

30. NIH halts clinical trial of hydroxychloroquine | National Institutes of Health (NIH). https://www.nih.gov/news-events/news-releases/nih-halts-clinical-trial-hydroxychloroquine.

31. Self, W. H. et al. Effect of hydroxychloroquine on clinical status at 14 days in hospitalized patients with COVID-19: a randomized clinical trial. JAMA 324, 2165–2176 (2020).

32. Sands, K. et al. No clinical benefit in mortality associated with hydroxychloroquine treatment in patients with COVID-19. International Journal of Infectious Diseases 104, 34–40 (2021).

33. Chivese, T. et al. Efficacy of chloroquine and hydroxychloroquine in treating COVID-19 infection: A meta-review of systematic reviews and an updated meta-analysis. Travel Med Infect Dis 43, 102135 (2021).

34. Kashour, Z. et al. Efficacy of chloroquine or hydroxychloroquine in COVID-19 patients: a systematic review and meta-analysis. Journal of Antimicrobial Chemotherapy 76, 30–42 (2021).

35. Budhathoki, P., Shrestha, D. B., Rawal, E. & Khadka, S. Corticosteroids in COVID-19: Is it Rational? A Systematic Review and Meta-Analysis. SN Compr Clin Med 2, 2600–2620 (2020).

36. van Paassen, J. et al. Corticosteroid use in COVID-19 patients: a systematic review and meta-analysis on clinical outcomes. Crit Care 24, 1–22 (2020).

37. Sterne, J. A. C. et al. Association between administration of systemic corticosteroids and mortality among critically ill patients with COVID-19: a meta-analysis. JAMA 324, 1330– 1341 (2020).

38. Group, T. R. C. Dexamethasone in Hospitalized Patients with Covid-19. 10.1056/NEJMoa2021436 384, 693–704 (2020).

39. Sterne, J. A. C. et al. RoB 2: a revised tool for assessing risk of bias in randomised trials. BMJ 366, (2019).

40. Falci, S. G. M. & Marques, L. S. CONSORT: when and how to use it. Dental Press J Orthod 20, 13 (2015).

41. Cuschieri, S. The STROBE guidelines. Saudi J Anaesth 13, S31 (2019).

