## Supplementary Materials for "Incorporating Preprints in Systematic Reviews: A Preliminary Study of a Novel Method for Rapid Evidence Synthesis"

### A. Data resources

#### A.1 Databases and Search Strategy:

For preprints, we systematically searched for studies of treatments for COVID-19. We searched the medRxiv, bioRxiv, arXiv, SSRN Electronic Journal, ChemRxiv, JMIR Preprints, and Research Square databases to September 03, 2020 using the following search terms: “COVID-19” or “SARS-CoV-2” and “clinical” and “treatment” and the names of potential treatments including^1^: chloroquine, hydroxychloroquine, azithromycin, carrimycin, convalescent plasma, ASC09, atazanavir, darunavir, danoprevir, lopinavir, ritonavir, stem cells, clazakizumab, olokizumab, sarilumab, siltuximab, sirukumab, tocilizumab, and remdesivir. For published articles, we used the same search terms and searched for meta-analysis studies on COVID-19 treatments indexed in PubMed.

#### A. 2 Eligibility Criteria:

Eligibility was restricted to studies of treatments or drug-related human experimental studies for COVID-19. Studies were excluded if the study type was in vitro or animal testing. Case reports, case series, observational studies with single or multiple arms, and randomized controlled trials (RCT) with any size of study population were eligible for inclusion. For preprints, we excluded literature reviews of COVID-19 treatments or those reporting meta-analysis results. Using this strategy, we identified 146 preprints posted on the preprint servers. As of September 03, 2020, 23 out of the 146 preprints have been published in a peer-reviewed journal. Of the 146 preprints, there were 7 preprints on hydroxychloroquine and 7 preprints on corticosteroids included in our case studies. In the published literature, we identified 6 published studies on hydroxychloroquine and 19 published studies on corticosteroids. Studies with both control (i.e., placebo or standard care) and treatment groups were eligible to be included.

#### A. 3 PRISMA Diagram:

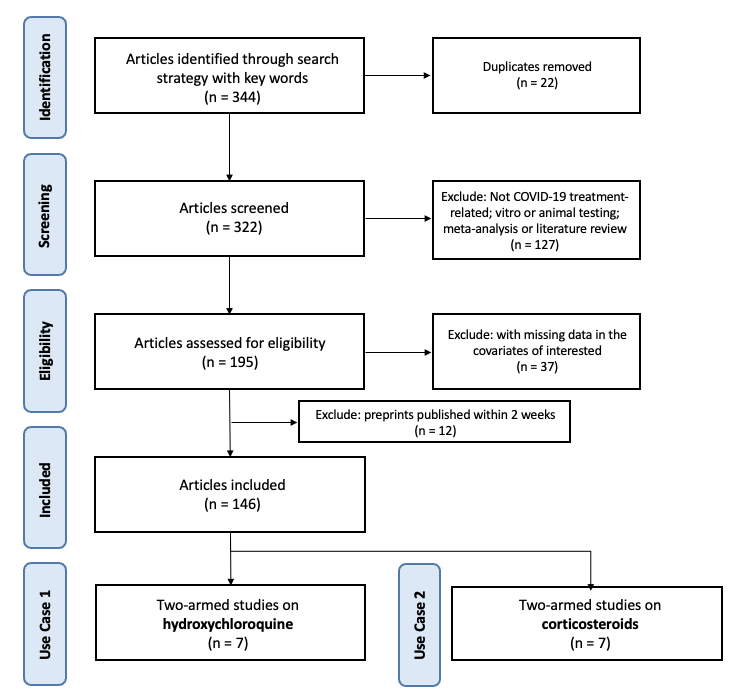

#### A. 4 Data extraction:

Data from the included manuscripts were extracted and organized in a systematic format. Specifically, for the case studies and survival cure model, we extracted various features from the eligible studies, including general information (title, authors’ names), publication/post date, population sizes, countries, single or multiple center studies, study types, number of arms, sizes of the arms, number of deaths in the arms, mortality rates, p-values of mortality rates, cured patients rates, participants’ median ages, number of males, preliminary results or not, and adjusted analysis for confounding variables or not. Other data of interest included citation counts through 09/03/2020, number of PDFs downloaded on the preprint servers through 09/03/2020, and h-index from Google Scholar of the last author as of 09/03/2020. This set of features is a set of the covariates extracted from the preprints, which has minimal missing values.

### B. Statistical method

Let $1-\pi(z)$ denote the probability that a manuscript will never be published depending on some study-level characteristics $z$ (i.e., $\pi(z)$ is the probability that a preprint to be published), and $S_{T}(t|x)$ be the probability of $T>t$ for those that will be published, which depends on features $x$. The mixture model assumes that the probability that a preprint has not been published by time $t$ is:

$S_{T}\left( t | x \right)=\pi\left( x \right)S\left( t | x \right)+\left( 1-\pi\left( x \right) \right)$	(1)

We use logistic regression with logit link to model $\pi\left( x \right)$ and proportional hazards (PH) regression to model $S\left( t | x \right)$, i.e., $\pi\left( x \right)=exp(x\gamma)/(1+exp(x\gamma))$ and $S\left( t | x \right)= S_{0}{(t)}^{\exp\left( x\beta\right)}$, where $\beta$ is the coefficient of the effects of $x$, and $S_{p}\left( t | x \right)= S_{0}{(t)}^{\exp\left( x\beta\right)}$, where $\beta$ is the coefficient of the effects of $x$ and $S_{0}(t)$ is the baseline survival function. The analysis is conducted using the R package “*smcure*”^22,23^.

For the published preprints, the confidence score is denoted as $\pi\left( x \right)$. For the not yet published ones, the confidence score should reflect the probability of eventual publication, given both study features and the fact that the study has not yet been published. Specifically, we define $u$ as the time between posting and analysis. Instead of using $\pi\left( x \right)=P\left( T<\infty\mid x \right)$ that only accounts for study features, we calculate the probability $P\left( T<\infty\mid T\geq u, x \right).$Under model (1), this probability (i.e., confidence score) can be expressed as $\left\{ S\left( u | x \right)-1+\pi(x) \right\}/S(u|x)$. The features in the logistic regression model $\pi\left( x \right)$ can be different or a subset of the variables in proportional hazard (PH) regression model.

For evidence synthesis, we included both published studies and preprints. Suppose there are $n$ total studies. We use $w_{i}$ to denote the weight for the $i$th study in the meta-analysis, $i=1,\ldots, n$. If the $i$th study is already published at the time of analysis, we set $w_{i}$to be 1. If the $i$th study is not published yet, its probability of never being published is estimated from the survival mixture model. The weight of the i-th study in the meta-analysis is set as $w_{i}=\left\{ S\left( u_{i} | x_{i} \right)-1+\pi(z_{i}) \right\}/S(u_{i}|x_{i})$ if it is not published. It's worth noting that when two studies with the same features remain unpublished at the time of meta-analysis, the earlier posted study is assigned a lower weight.

Based on the $w_{i},$which characterizes the chance of publication for the i-th study, we propose the multiple imputation procedure for evidence synthesis. For each study, we impute the status of publication $s_{i}$, where $s_{i}$ = 1 indicates studies that will be published, and $s_{i}$ = 0 indicates a study that will never be published, from a Bernoulli distribution with probability $w_{i}.$ We then conduct a random-effects meta-analysis using all the studies with $s_{i}$ = 1. We can estimate an effect size, and standard error of the estimated effect size, as well as the heterogeneity variance $\tau^{2}$. By repeating the above imputation-estimation process multiple times, we can obtain a final estimate for the overall effect size by taking the average of the estimated effect sizes for all imputations, as well as a final estimate for the heterogeneity variance. In the case studies, this imputation process was repeated 500 times.

### C. Sub-analysis with RCTs only

For both case studies, we also conducted the analysis on the collected RCTs only with the proposed two-stage procedure. For the case study on hydroxychloroquine, if we limited our analysis to RCTs, the relative risk (RR) of 5 published articles is 1.23 (95% CI: 0.56, 2.73) and the RR value with the proposed method on 6 RCTs (5 published and 1 preprint) is 1.08 (95% CI: 0.56, 2.28). For corticosteroids, the RR of 8 published articles is 0.84 (95% CI: 0.74; 0.96) and the RR value with the proposed method on 9 RCTs (8 published and 1 preprint) is 0.84 (95% CI: 0·69, 1.03).

1	COVID Trial Explorer | Tableau Public. https://public.tableau.com/app/profile/marinamarin/viz/covidTrials/COVID-19ClinicalTrialsExplorer (accessed Sept 6, 2021).

### D. Examples and Guidance on Choice of Strategies

The choice between Strategy III and Strategy IV depends on the practical context. Strategy III offers simplicity and interpretability by applying a binary inclusion/exclusion rule based on a confidence score. However, this approach may result in the exclusion of potentially valuable information from preprints with lower confidence scores. In contrast, Strategy IV retains the full spectrum of preprints and adjusts their influence proportionally to their estimated likelihood of publication. This method is more inclusive and statistically principled. We have provided some examples and guidance on how to choose the strategy in practice below.

- If a substantial number of preprints have confidence scores clustered near the threshold (e.g., many scores in the range of 0.45–0.55), small perturbations in CS or threshold choice can lead to discrete shifts in inclusion/exclusion under Strategy III.
  - For instance:
    - Suppose 10 preprints have CS values of 0.48–0.52. Under Strategy III with a threshold of 0.5, half of them may be included, while minor changes in the model or data could flip the inclusion of all 10.
    - This threshold sensitivity can lead to instability in the meta-analytic estimate, especially when preprints comprise a large proportion of the total evidence.
  - In contrast, Strategy IV would smoothly down-weight preprints with mid-range CS values, mitigating the impact of threshold arbitrariness. Therefore, Strategy IV may be more robust when:
    - CS values fall within an ambiguous range (e.g., 0.4–0.6),
    - The number of preprints is large relative to published studies
- If most preprints have very high confidence scores (e.g., >0.9), the threshold approach effectively mimics the inclusion of peer-reviewed articles, making Strategy III nearly equivalent to Strategy IV in terms of results but simpler to interpret and apply.
  - For example:
    - If 8 out of 10 preprints have CS > 0.95, the effect estimate from Strategy III (thresholding) and Strategy IV (weighting) will converge, since nearly all studies receive full weight in both.
    - In such cases, Strategy III may be preferred for its transparency and ease of communication, particularly in policy-facing or clinical decision-making contexts.
